# Mortality Analysis of COVID-19 Confirmed Cases in Pakistan

**DOI:** 10.1101/2020.06.07.20121939

**Authors:** Ambreen Chaudhry, Aamer Ikram, Mirza Amir Baig, Muhammad Salman, Tamkeen Ghafoor, Hussain Zakir, Mumtaz Ali Khan, Jamil Ahmed Ansari, Asif Syed, Wasif Javed, Ehsan Larik, Muhammad Mohsan, Naveed Masood, Zeeshan Iqbal, Khurram Akram

## Abstract

**Introduction:** COVID-19, a novel disease, appeared in December 2019 in China and rapidly spread across the world. Till second week of April 2020, high incidence (2.9/100,000) and cases fatality rates (1.7%)was observed in Pakistan.

This study was conducted to determine temporal and spatial distribution of first 100 deaths attributed to COVID-19 in Pakistan and their associated demographic factors.

**Method:** We conducted a descriptive epidemiological analysis of first 100 deaths reported among RT-PCR confirmed COVID-19 cases. Demographic, epidemiological and risk factors information was obtained associated comorbidities and clinical signs and symptoms were recorded and frequencies were determined.

**Results:** A total of 100 mortalities with overall Case Fatality Rate 1.67% (CFR) were analysed. Median age of patients was 64.5 years (IQR: 54–70) with 75% (n = 75) Males. Among all deaths reported, 71 (71%) cases had one or more documented comorbidities at the time of diagnosis. Most frequently reported co-morbidities were; hypertension (67 %), followed by Diabetes Mellitus 945%) and Ischemic Heart Diseases (27%). First death was reported on 18 March 2020 and the most frequent presenting symptoms were shortness of breath (87%) and fever (79%). Median duration of illness was eight days (IQR: 4–11 days), median delay reaching hospital to seek health care was three days (IQR: 0–6 days) while median duration of hospital stay was also three days (IQR: 1–7 days). Among all reported deaths, 62% were attributed to local transmission as these cases had no history of international travel. The most affected age group was 60–69 years while no death reported in age group below 20 years.

**Conclusion:** High CFR among old age group and its association with co-morbidities (chronic disease) suggests targeted interventions such as social distancing and strict quarantine measure for elderly and morbid people. Comparative studies among deaths and recovered patients are recommended to explore further disease dynamics.

## Introduction

In December 2019, several cases of pneumonia of unknown causes were reported from Hubei, China later named as severe acute respiratory syndrome coronavirus-2 (SARS-CoV-2). The international Committee on Taxonomy of Viruses renamed the virus as acute respiratory syndrome coronoavirus-2 (SARS-CoV-2).(1) The World Health Organization (WHO) announced the epidemic caused by SARS-CoV-2 as coronavirus disease 2019 (COVID-19). In subsequent days, world has seen a rapid spread across international borders and high rates of morbidity and mortality. (2) Considerable efforts has been made to understand the mechanism of disease arose as severe respiratory disease with 89.1% nucleotide similarity to a group of SARS-like coronavirus found in bats in China (3). Many developed countries with strong health infrastructure are facing high mortality hit. Case Fatality estimation has suggested a range of 0.25%-3.0% with highest 3.5 in China alone at its earliest(4). However, a correct estimation of the disease is still needed as the situation is changing rapidly. Some researcher suggested an unadjusted range of 4.4% to 4.8% considering rest an underestimation(5). Initial estimates among Novel Corona Infected Pneumonia (NCIP) suggested a human to human spread in symptomatic as well as asymptomatic(6). No significant difference in viral load of symptomatic and asymptomatic people (7) lead to the outcome of global spread as well as an underestimated mortality rate. (8).

Morbidity and mortality in developed countries has been documented high, attributed to having big proportion of aging population as compared to china(9). As of today, we know that virus is affecting badly the extreme ages and those with co-morbidities. (10).

Analysis of fatal cases in China has shown high rates in patients with co-morbidities of Ischemic Heart Diseases, Hypertension, and Diabetes. However, a higher risk in pregnant women has not been established so far.(11) Literature shows a death rate among hospitalized individuals was 15% with mean period of 14days from onset of symptoms to deaths of patients. (12).

Since first confirmed case reported on 26^th^ Feb and first death reported on 12^th^ March, 2020, number of cases are increasing exponentially and so is the case fatality rates. (13) In the light of rapidly changing epidemic situation, a rapid and ongoing epidemiological analysis of morbidities and mortalities was pertinent for timely and robust public health responses.

Our objective was to investigate the characteristics of Patients died of novel corona disease, their spatial and temporal distribution and risk factors associated with them.

## Methodology

A retrospective record review was conducted at National Institute of Health (NIH) Islamabad, to analysed first 100 deaths reported and recorded with the National Emergency Operation Centre of NIH (NEOC), among COVID-19 cases, confirmed through RT-PCR. Demographic, epidemiological and risk factors information was obtained and their history of travel was probed.

Overall case fatality rate was calculated and its spatial and temporal trends were determined. Associated comorbidities and clinical signs and symptoms were recorded and their frequencies were calculated.

Study was approved by the National Institute of health’s ethical review board and was exempted from written informed consent by the same. All the enrolled mortalities were diagnosed as COVID-19 positive in accordance with national guidelines using Real time PCR techniques at designated testing centres across Pakistan. Along with patients’ available records review, verbal autopsies were carried out where appropriate, to gather maximum information as possible. All identifiers were removed before entering patient’s data. Basic descriptive analysis was conducted using Epi Info® 7.

## Results

A total of 100 mortalities (Overall CFR-1.67%) were analysed. Incidence and mortalities showed an exponential trend over time (Fig: 1).

**Figure 1:**
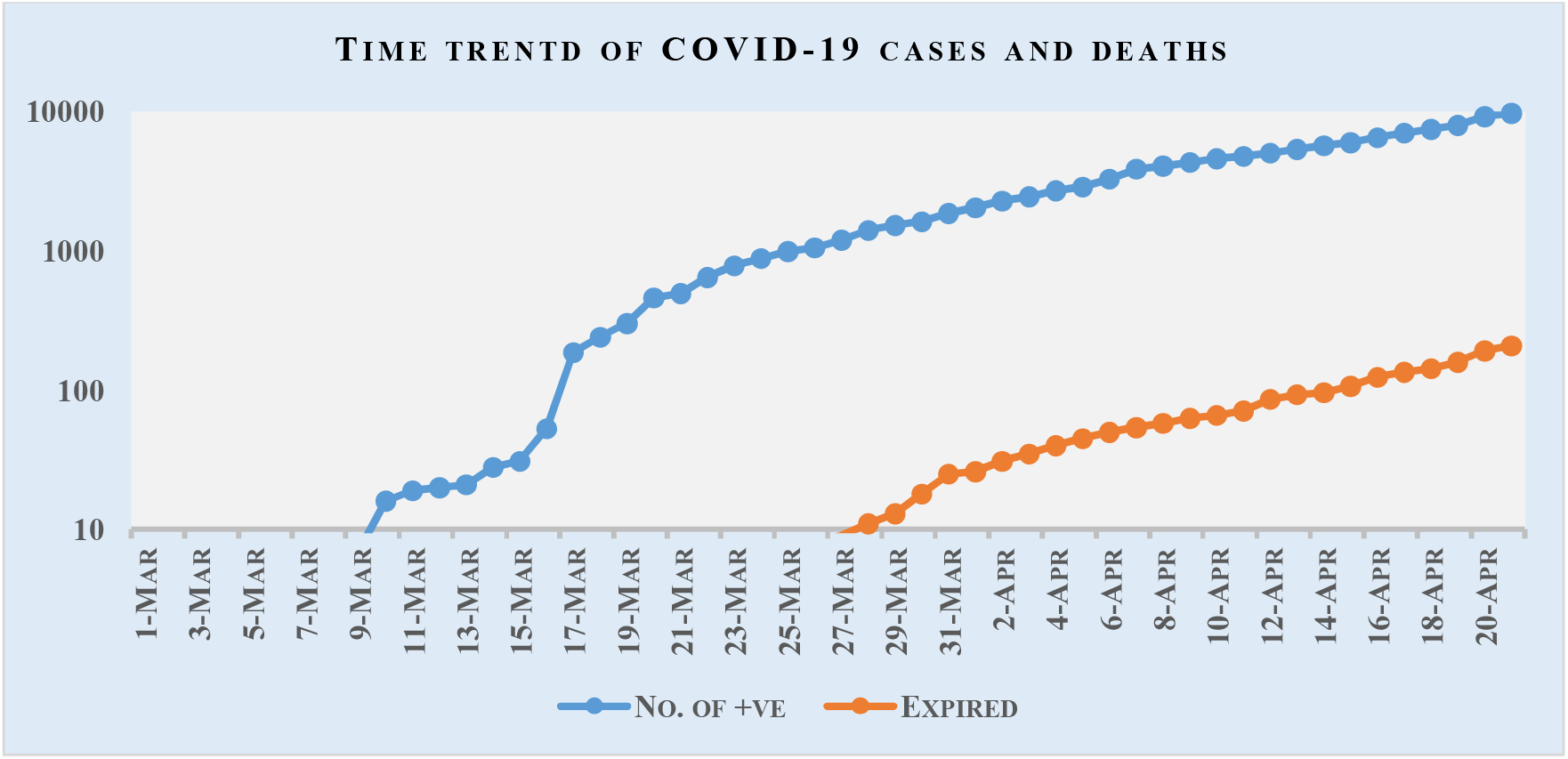
Incidence and mortality rates of COVID-19 in Pakistan

Median age of patients was 64.5 years with a range of 22–84years. 75% (n = 75) cases were Males. Among all deaths reported 71 (71%) cases had a documented co-morbidity at the time of presentation. The most frequently reported co-morbidity among all deaths was hypertension (67%) followed by Diabetes Mellitus (45%) and Ischemic Heart Diseases (27%). First death was reported on 18^th^ of March and most frequent presenting signs/symptoms were shortness of breath (87%) and fever (79%). Median duration of illness was eight days (range: 0–30 days), median delay in reaching to hospital was three days (range: 0–16) while median duration of hospital stay was three days (range: 0–28 days). Among all deaths reported, 62% were attributed to local transmission and had no history of international travel (table 1).

**Table 1:**
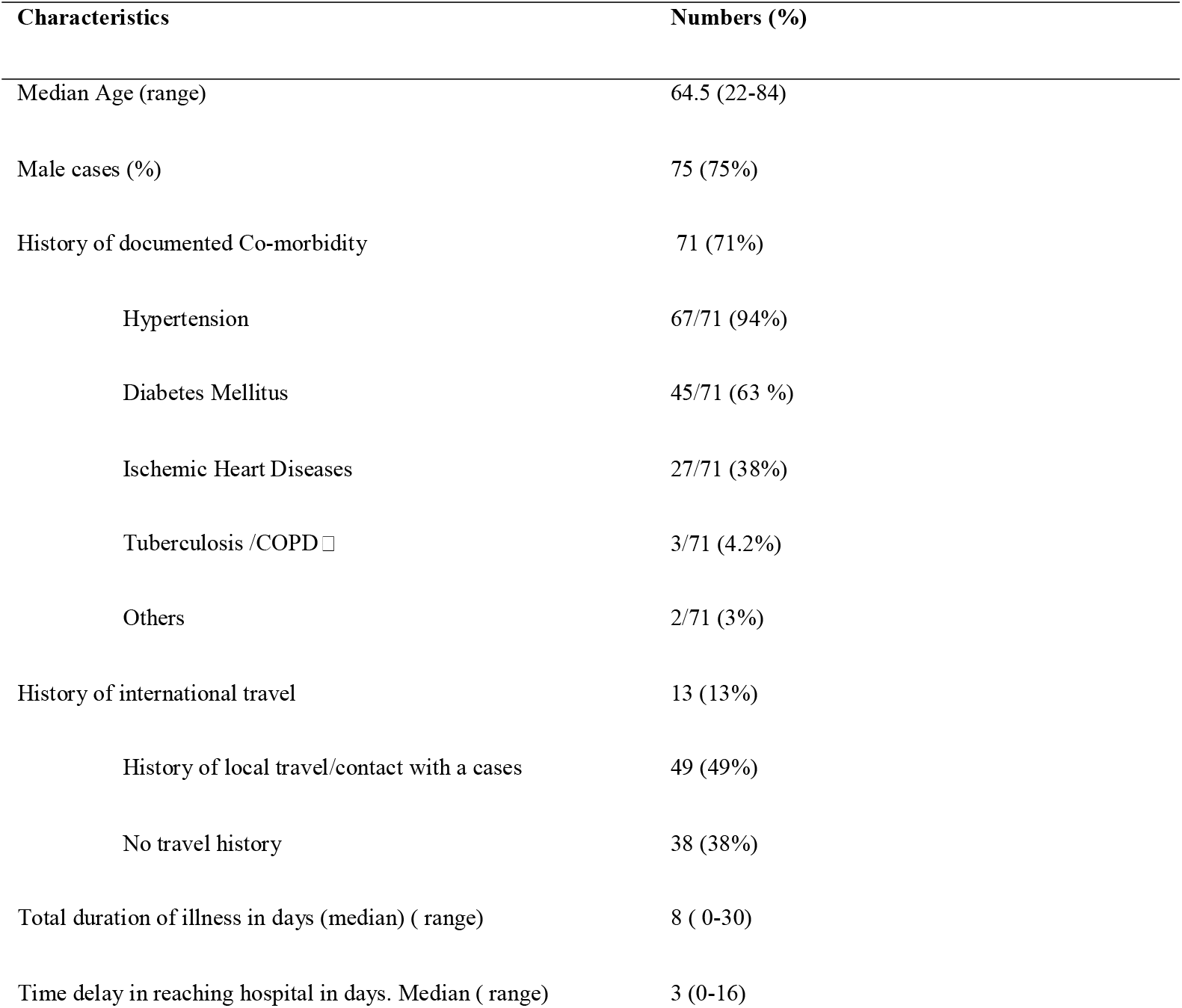
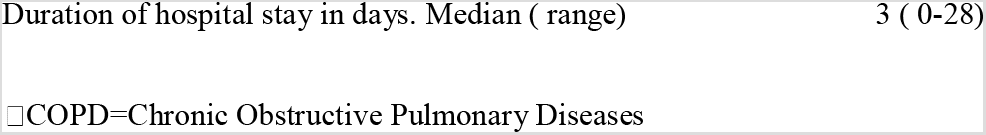
Description of epidemiological characteristics of first 100 deaths attributed to CVID-19 in Pakistan.

| Characteristics | Numbers (%) |
| --- | --- |
| Median Age (range) | 64.5 (22-84) |
| Male cases (%) | 75 (75%) |
| History of documented Co-morbidity | 71 (71%) |
| Hypertension | 67/71 (94%) |
| Diabetes Mellitus | 45/71 (63 %) |
| Ischemic Heart Diseases | 27/71 (38%) |
| Tuberculosis /COPD | 3/71 (4.2%) |
| Others | 2/71 (3%) |
| History of international travel | 13 (13%) |
| History of local travel/contact with a cases | 49 (49%) |
| No travel history | 38 (38%) |
| Total duration of illness in days (median) ( range) | 8 ( 0-30) |
| Time delay in reaching hospital in days. Median ( range) | 3 (0-16) |

□ COPD=Chronic Obstructive Pulmonary Diseases

Among all age groups, 60–69 years were most affected while no death reported in below 20 years age groups (table 2)

**Table 2:**
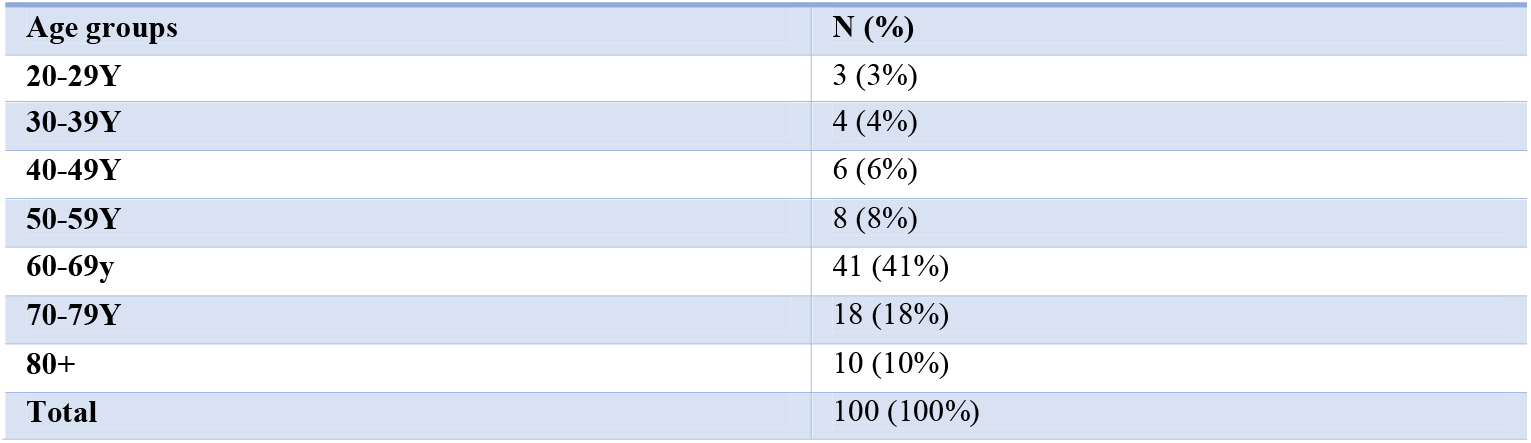
Age distribution of first 100 deaths attributed to COVID.

Highest case fatality rate was observed in Khyber Pakhtoon khwa (KP) province (4.39%), while no deaths was reported from Azad Jammu and Kashmir region (fig 2)

**Figure 2:**
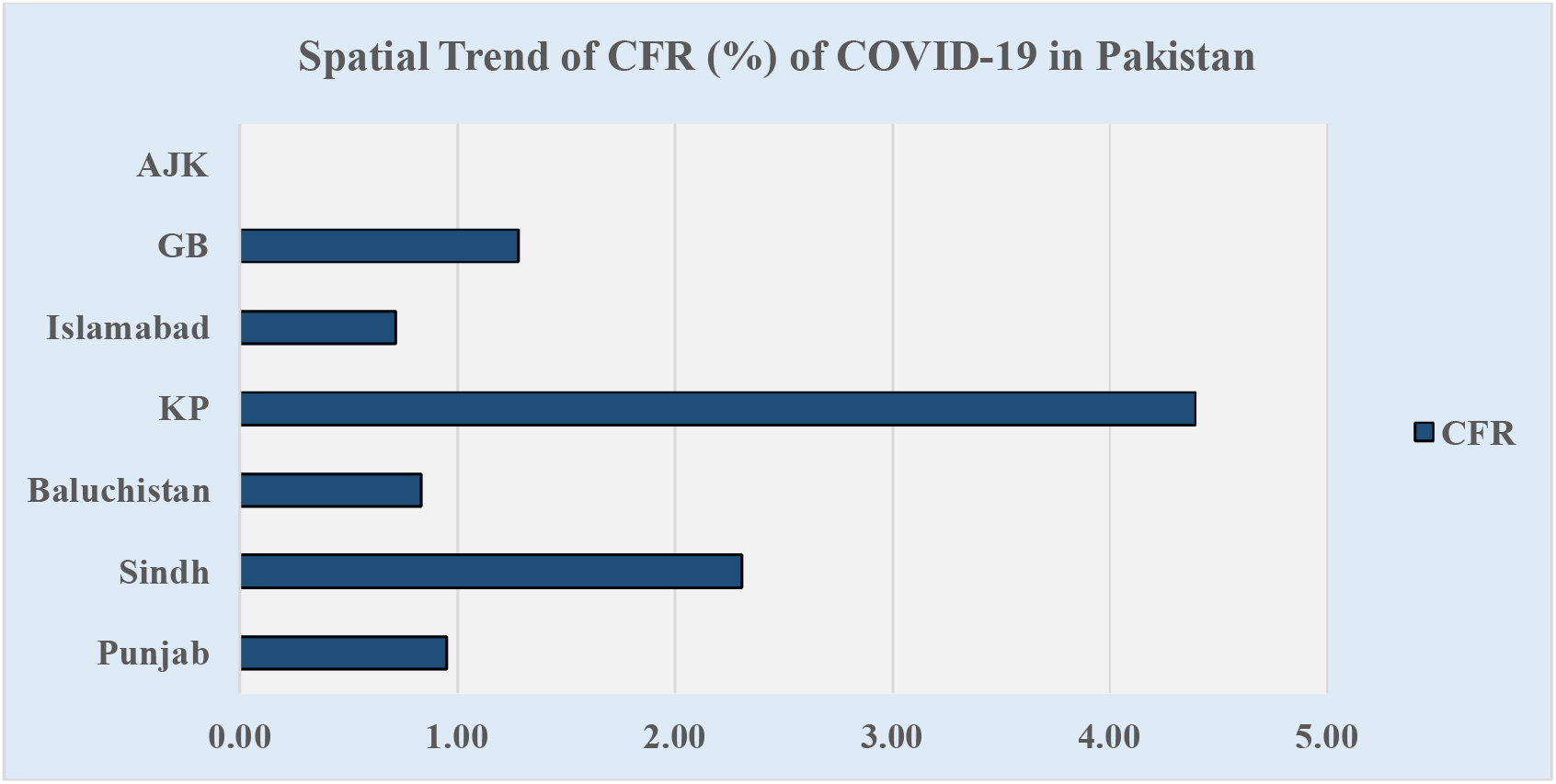
Geographical distribution of case fatality rate in Pakistan

## Discussion

During the early phase of outbreak, reported cases were mostly travellers from other countries, mainly Iran. Testing for COVID-19 was done for symptomatic as well as asymptomatic people who had an international travel history. First death was reported on 18^th^ March with gradual increase in CFR to 1.67% till first 100 deaths were observed, which is well below the level estimated by some researchers. (5) Initial phase of epidemic shows more cases having international travel history (14) which later on lead to local transmission. Early quarantine of travellers, prompt adoption of social distancing as well as national level lockdown policies might have played their roles in decelerating the epidemic curve. Our results showed that age above 60 years is the most vulnerable age group as depicted in other studies. (15) Children and young age groups are seem to be protected while middle age group having highest community exposure are mildly affected. An increased CFR has been observed with increasing age.

Cases presented with a range of signs and symptoms. Fever, cough and breath shortness are the most reported signs and symptoms. However, severe acute respiratory syndrome seems the hall mark of mortality. Patients of 60 years and above presented with a rapidly progressing disease (16) along with fever, sore throat and cough as presented in other countries. (10) Patients above 60 years are dependent group of our society so are less than 12 years and usually have little community exposure as compared to rest of the population. Since results showed no death reported below 20 years age group, high CFR among 60+ patients is relatable to their low immunity and co-morbidities. Our results showed very short span of illness and even shorter duration of hospitalization overtimes. Since the prevention measures were being observed all acorns the country, access to hospital and health seeking behaviour of community played a major role in cases registration. Comorbidities like Ischemic heart diseases, diabetes and hypertension seems to play a critical role in disease progressions as determined by some other authors.(17)

Highest CFR was observed in KP province (4.39%) followed by Sindh province (2.31%). Karachi city, the most populous metropolitan and hence most affected by epidemic in Sindh province, observed early preventive measures as compared to rest of country. There is direct association between social distancing and disease transmission rate. (18) Hence a rise in CFR in KP and low in GB are directly relatable to their social distancing policies.

## Conclusion

We concluded that a high fatality among elderly population and its association with co-morbidities gives us a chance to taper our intervention. Standard Operating Procedures for quarantine and isolation of elderly should be revised. Keeping our traditions and cultural norms regarding respect and care of elderly, home quarantine measures should be robust in big cities like Karachi. Male population is more prone linking their social activities in a male dominant society. Analysis of first 100 COVID-19 related deaths is a cue to assess the outcome of measures taken so far. All provinces should take all necessary measures to protect their vulnerable population to lower the CFR further down.

## Limitation

Though this descriptive analysis has shown a trace of epidemiological measures, a comparative study is suggested to assess the factors associated with death as compared to recovery of the COVID-19 patients. Initial mortalities rates may vary from subsequent mortality patterns where local transmission will be high.

## Ethical considerations

Patients were not directly involved therefore informed written consent was not obtained obtained.

## Data Availability

Data is available on request, subject to the approval of National Institute of Health of Pakistan

## Acknowledgment

We are thankful to Technical support Officers of District Surveillance and Response Units of FELTP Pakistan for their technical support.

## Author’s Contributions

AC, KA, FK conceived the idea, analysed data and wrote the manuscript. MAK, MAB, ZH and AI provided critically reviewed.

